# Prevalence and correlates of joint pain on physical, mental, and social health in women living with and without HIV

**DOI:** 10.64898/2025.12.02.25341477

**Authors:** Ryan D. Ross, Elizabeth Daubert, Stacey M. Cahoon, Ruby Hernandez, Darlene Johnson, Tracey E. Wilson, Audrey L. French, Mardge H. Cohen, Kathleen M. Weber

**Affiliations:** Departments of Anatomy & Cell Biology and Microbial Pathogens & Immunity, Rush University Medical Center, Chicago, IL; Hektoen Institute of Medicine, Chicago IL, USA; Department of Community Health Services at the State University of New York Downstate Medical Center; Department of Medicine, University of Illinois Chicago, Chicago IL, USA; Department of Medicine, Cook County

**Keywords:** Pain, Joint Pain, Depression, Anxiety, Frailty, Loneliness

## Abstract

**Introduction:** Joint pain is amongst the most common and disabling form of chronic pain globally and is more frequently reported by women than men. Whether joint pain is more prevalent among women with versus without HIV (WWH/WWoH) and differentially contributes to frailty phenotypes and physical, mental, and social functional impairment is unknown.

**Methods:** The Common Data Elements (CDE) Pain, Enjoyment, and General Activity (PEG) instrument was used to assess and compare joint pain in Chicago midlife WWH versus WWoH and its association with key outcomes (frailty, depressive symptoms, anxiety, and loneliness) using stepwise multivariable models.

**Results:** PEG was assessed in 179 WWH and 81 WWoH. Overall, 42% of women reported moderate/severe pain, which was more commonly reported in WWH than WWoH (44.1 v. 38.3%) but did not reach statistical significance. Women with moderate/severe joint pain, compared to those with no/mild joint pain (58%) were more likely to be pre-frail/frail (aOR 3.05; CI: 1.65-5.63) and report greater depressive symptomology (aOR 2.42; CI: 1.31-4.47), anxiety (aOR 3.38; CI: 1.52-7.43), and loneliness (aOR 1.84; CI: 1.02-3.29).

**Conclusions:** Joint pain is highly prevalent in mid-life WWH and WWoH and is associated with greater physical, mental, and social health burdens. Incorporating the PEG as a screening tool in clinical practice may help identify women who would benefit most from joint pain interventions to enhance functional capacity.

## Introduction

Chronic pain is a highly prevalent symptom reported by people with HIV (PWH) (1), and most commonly due to degenerative musculoskeletal (MSK) disorders such as osteoarthritis that contribute to joint and low back pain (2). MSK disorders are more common in women especially after menopause and with increasing age and body mass index (BMI) (3, 4). Importantly, with effective therapy, the average age of women with HIV (WWH) in the US is greater than 50 years and more WWH are entering menopause (5), which will likely increase the burden of joint pain. Currently, the impact of joint pain remains unclear as much of the chronic pain data in HIV populations published to date does not isolate joint pain specifically. Further, lack of comparisons to demographically-similar women without HIV (WWoH) limit understanding about the direct effects of HIV and its treatment on MSK pain (6).

Joint pain can negatively impact quality of life, influencing physical, mental, and social health. Osteoarthritis pain, for example, is associated with impaired physical functioning (7) and greater anxiety and depression (8). Similar effects have been noted in people with low back pain (9, 10). People reporting greater MSK-related pain also report higher levels of loneliness (11). Whether chronic joint pain contributes to similar adverse effects in WWH is currently unknown. Therefore, the current study sought to investigate the prevalence and the physical, mental, and social health correlates of self-reported joint pain in midlife WWH and demographically similar WWoH using a common data elements compliant pain instrument.

## Methods

### Study Population

The Multicenter AIDS Cohort Study (MACS)/Women’s Interagency HIV Study (WIHS) Combined Cohort Study (MWCCS) is an ongoing, multicenter, observational study of the treated history of HIV. Informed written consent was obtained from participants prior to study enrollment and all analysis was approved by Rush University Medical Centers Institutional Review Board. Using structured interviews and standardized protocols, staff collected data on clinical, sociodemographic, behavioral, and environmental factors as well as physical parameters and biologic specimens. Study methods and national cohort characteristics have been described previously (12). The Chicago Cook County site of the MWCCS enrolled women with HIV (WWH) and socio-demographically similar women without HIV (WWoH) in multiple waves beginning in 1994. During the most recent annual research visit (*October 1, 2023 – September 30, 2024)*, a total of 260 (83% of eligible participants) women (179 WWH; 81 WWoH), over the age of 40 years, completed a joint pain assessment during their in-person research visit. When compared to women enrolled in the Chicago Cook County site eligible but not included in the joint pain assessment, pain respondents were less likely to be WWH, more likely to be Black, and more likely to have a high school education or less (Supplemental Table 1).

### Joint Pain Assessment

Joint pain was evaluated using the Pain, Enjoyment of life, and General activity (PEG), a brief validated three-item scale derived from the Brief Pain Inventory (BPI) to assess pain intensity and interference with general activity and enjoyment of life (13). The PEG was developed in 2009 as a shorter and more easily scored pain common data element (CDE) compliant measure, which has been validated to be reliable through comparisons to other commonly used pain instruments, including the BPI, the Chronic Pain Grade questionnaire (CPG), and the Short-Form 36-item questionnaire (SF-36) (13).

The standard PEG form includes three items to assess pain level and interference for any joint. We employed two versions of an expanded PEG instrument over the course of the sub-study. The initial expanded version was administered to 125 women between October 2023 and February 2024, while a shorter version was administered to 135 women between February 2024 to October 2024. The expanded version asked women to identify the number that best describes their joint pain for each joint area (knees, hips, and spine) on average and how that anatomic specific joint pain interferes with enjoyment of life and general activity during the past week. Therefore, a total of 9 questions regarding pain were asked – three for each of the three joint regions. An additional question was included to ascertain the specific joint representing the *<u>most</u>* pain as right versus left for knee and hip and upper versus lower for spine pain (attached as Supplemental Form 1).

The PEG was subsequently shortened after determining the longer form did not increase the ascertainment of usable data. We subsequently shortened our form to the original PEG format, three questions regarding pain in any joint, while still ascertaining the *<u>most</u>*painful joint along with capturing any additional painful joints not included in the combined query (attached as Supplemental Form 2). Each of the three severity and interference questions used the same 0-10 numerical rating scale with 0 being the least and 10 being the worst pain. A summary PEG score was calculated as the average of the three scaled severity and interference responses. For the expanded version of the PEG form, the summary PEG score was calculated using responses from the most painful joint. No/mild pain was defined as a score from 0 to <4, moderate pain as ≥4 to <7, and severe as ≥7 to 10.

### Fried Frailty Phenotype (FFP)

Frailty was categorized using the five components of the FFP. 1) Impaired mobility or slow walk speed was defined as ≥7 seconds for women ≤159 cm or ≥6 seconds for women >159 cm tall on the timed 4-meter walk. 2) Reduced grip strength was defined as the lowest 20% according to BMI and sex/gender, averaged across three dynamometer attempts with the dominant hand. 3) Heath-related physical limitations were determined by the response to the question: “How much (if at all) has your health limited you in the kinds or amounts of vigorous activities that you can do, like lifting heavy objects, running or participating in strenuous sports?” A response of “limited a lot” was considered positive for physical limitations. 4) Exhaustion was determined from the question of “during the past 4 weeks, have you been unable to do certain kinds or amounts of work, housework, schoolwork, or caring for children because of your health”, a response of “All of the time” or “Some of the time” was considered positive for exhaustion. 5) Finally, unintentional weight loss was defined as self-reported unintentional weight loss of ≥10 pounds in the previous year. Participants that met the frailty threshold for ≥3 FFP components were defined as frail; those meeting 1-2 FFP components were defined as pre-frail, and if no frailty components were met, women were categorized as robust.

### Depressive Symptom Burden

The 10-item version of the Center for Epidemiological Studies-Depression (CES-D-10) was used to assess the frequency in days of depressive symptoms over the past week on a four-point scale (rarely, some, occasionally, most). After reverse scoring the two positive mood items, all item responses were summed to obtain a total score ranging from 0-30 with higher scores indicative of greater depressive symptom burden. The total CES-D-10 score was dichotomized using a cutoff of ≥10 to represent presence of clinically significant depressive symptoms (14, 15).

### Anxiety Symptom Burden

Anxiety was assessed using the anxiety severity module of the Computerized Adaptive Testing-Mental Health (CAT-MH), termed the CAT-ANX. The CAT-ANX score was transformed to a 0 to 100 scale. Thresholds were defined empirically, with “no anxiety” defined as a CAT-ANX score of 35, mild anxiety as scores from 35 to 50, moderate anxiety as scores 50 to 65, and severe anxiety as scores 65 (16).

### Loneliness

Loneliness was assessed using the 3-item Loneliness Brief Form (17). Women were asked 1) “how often do you feel isolated from others?”, 2) “how often do you feel left out?”, and 3) “how often do you feel lack of companionship?”. The responses were scaled as “hardly ever” = 1, “some of the time” = 2, and “often” = 3. The results were summed, with the loneliness threshold set to a composite score ≥ 6{Friedman, 2021 #1591}.

### Additional Study Variables

Additional demographic variables included age in years, race/ethnicity (non-Hispanic Black, Hispanic, other, and non-Hispanic White), highest education attainment (less than high school, high school, or more than high school), partner status, and annual household income (≤$18,000 vs. □$18,000). Stages of Reproductive Aging Working Group (STRAW+10) criteria were used to categorize women as pre-menopause, perimenopause, or post-menopausal (18). Body mass index (BMI) was calculated using directly measured height and weight recorded at the study visit (kg/m^2^).

Lifestyle variables included cigarette use (current vs. former/never), alcohol use (none vs. ≤7 drinks/week vs. >7 drinks/week), marijuana use (current vs. former/never), crack/cocaine/heroin use (current vs. former/never).

Additional variables of interest in WWH included CD4+ T-cell count (CD4 cells/mm^3^), HIV viral load (categorized as detectable vs. undetectable with a threshold of 20 copies/mL), current use of ART, and total years since reported ART initiation.

### Statistical Analysis

Data was compared for normality and presented as means with standard deviations (SD), if normally distributed, and median with interquartile ranges (IQR), if not normally distributed. Demographic and lifestyle variables and self-reported joint pain were compared between WWH and WWoH using chi-square tests or Fisher’s exact when categorical groups had fewer than 5 women. Demographics, physical, mental, and social health indices were compared according to HIV-serostatus group using chi-square or Fisher’s exact tests.

Logistic regression models were constructed to investigate the associations between binary joint pain (v between participant characteristics and each of the four outcome variables. Participant characteristics with p-values <0.2 were used in subsequent stepwise multivariable (MV) models, with age and joint pain forced into each model and HIV-serostatus forced into the models that included the full study sample. Two separate stepwise MV models were run with the full study sample and then with WWH only.

## Results

### Women characteristics

A total of 260 Chicago MWCCS women over age 40 years completed the PEG survey (179 WWH and 81 WWoH). The mean age was 57.1 ± 8.6 years (Table 1). Most women identify as Black (69%) and late peri/postmenopausal (82%). Roughly 40% of women were current smokers, 11% reported consuming >7 drinks per week, 29% use marijuana and 29% have less than a high school education. WWoH were recruited into the parent study based on report of HIV risk behaviors and therefore, more likely to use crack, cocaine, or heroin (26% vs. 15% in WWoH vs. WWH, p=0.027). WWH reported using ART (97%), with a mean of ∼18 years of ART exposure, ∼76% had undetectable viral loads, with a median current CD4+ cell count of 761 cells/mL, and 77% had a CD4+ count >500 cells/mL

**Table 1:**
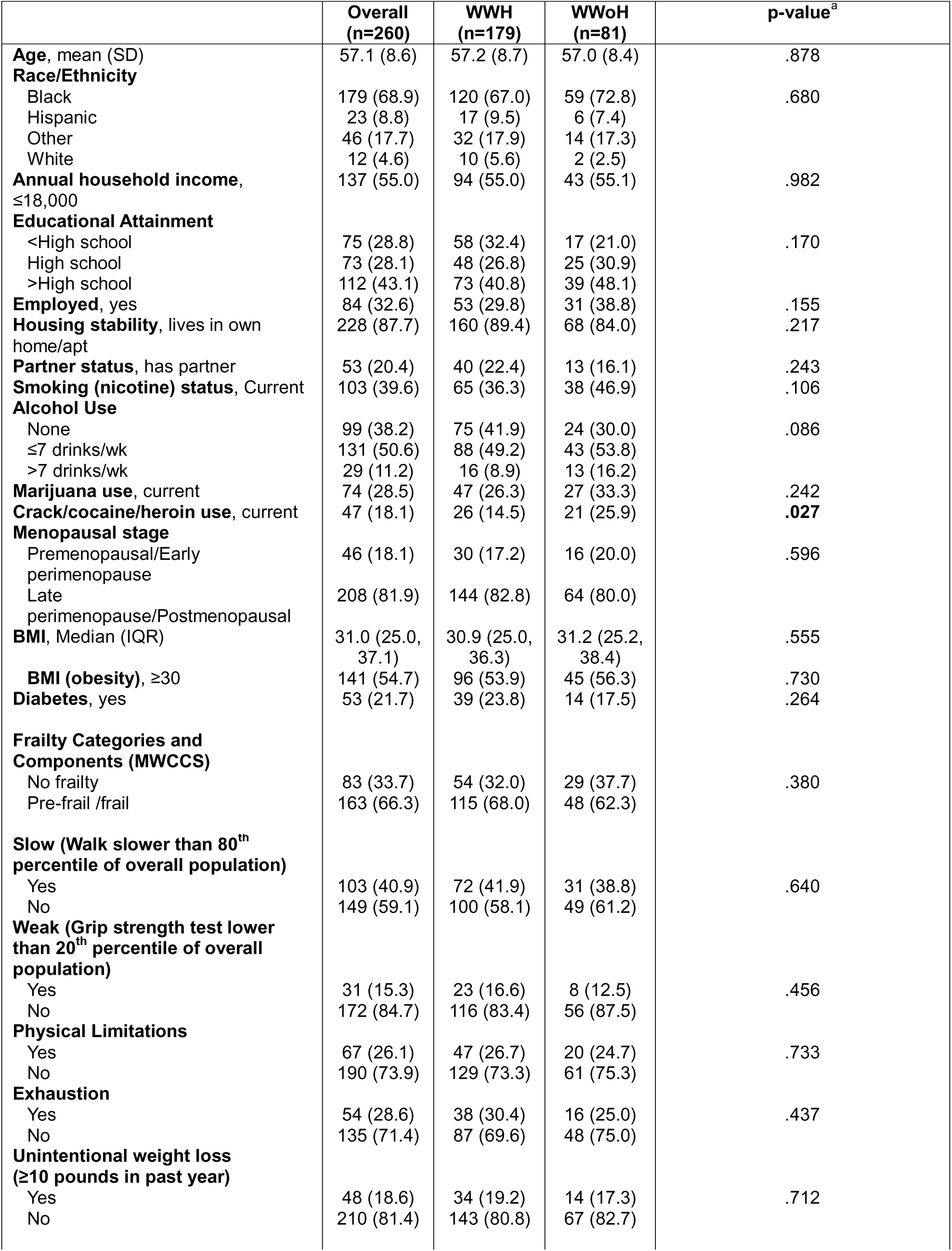
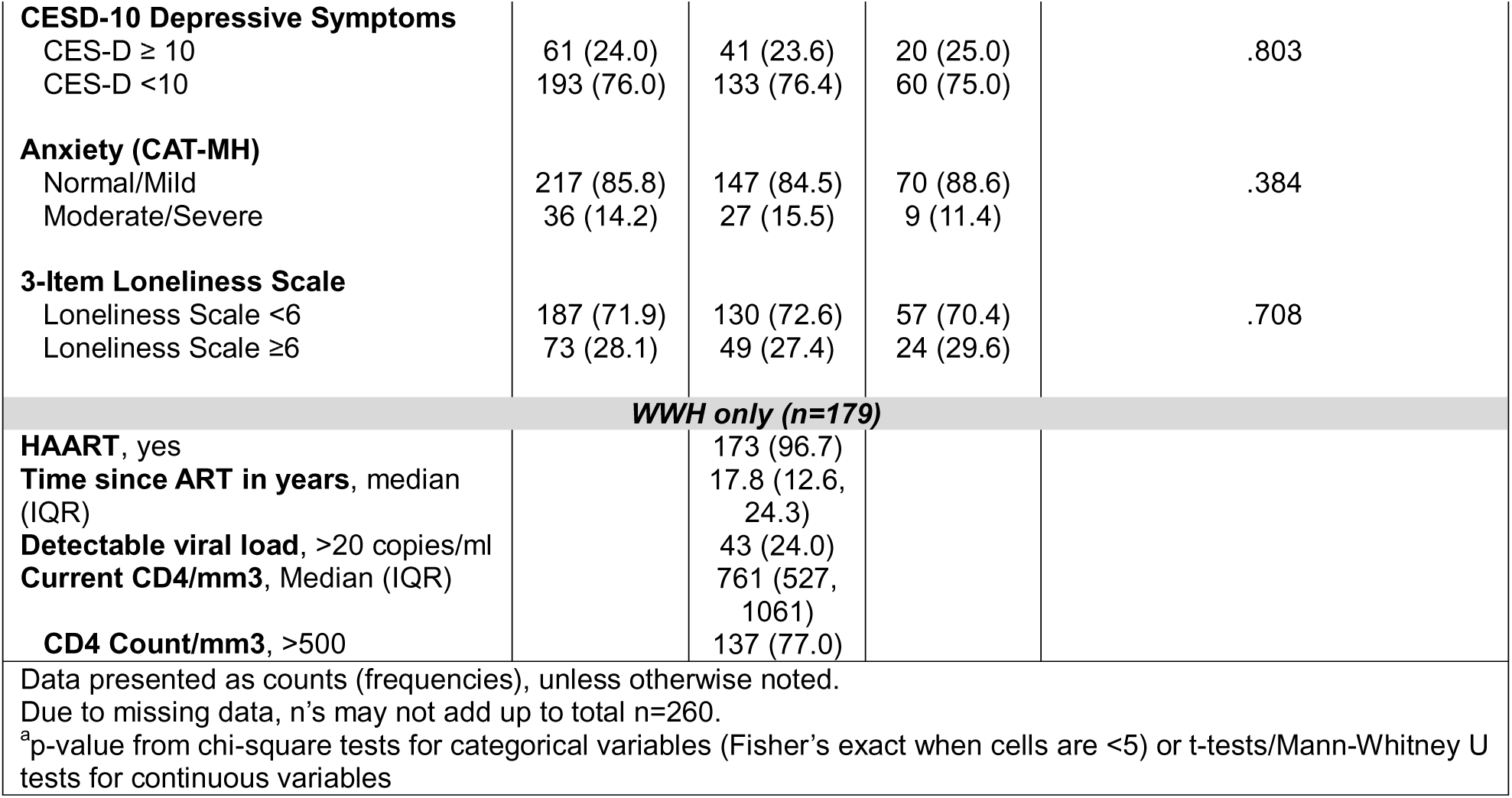
Participant characteristics.

|  | **Overall**<br>**(n=260)** | **WWH**<br>**(n=179)** | **WWoH**<br>**(n=81)** | **p-value<sup>a</sup>** |
| --- | --- | --- | --- | --- |
| **Age**, mean (SD) | 57.1 (8.6) | 57.2 (8.7) | 57.0 (8.4) | .878 |
| **Race/Ethnicity** |  |  |  |  |
| Black | 179 (68.9) | 120 (67.0) | 59 (72.8) | .680 |
| Hispanic | 23 (8.8) | 17 (9.5) | 6 (7.4) |  |
| Other | 46 (17.7) | 32 (17.9) | 14 (17.3) |  |
| White | 12 (4.6) | 10 (5.6) | 2 (2.5) |  |
| **Annual household income**,<br>≤18,000 | 137 (55.0) | 94 (55.0) | 43 (55.1) | .982 |
| **Educational Attainment** |  |  |  |  |
| <High school | 75 (28.8) | 58 (32.4) | 17 (21.0) | .170 |
| High school | 73 (28.1) | 48 (26.8) | 25 (30.9) |  |
| >High school | 112 (43.1) | 73 (40.8) | 39 (48.1) |  |
| **Employed**, yes | 84 (32.6) | 53 (29.8) | 31 (38.8) | .155 |
| **Housing stability**, lives in own<br>home/apt | 228 (87.7) | 160 (89.4) | 68 (84.0) | .217 |
| **Partner status**, has partner | 53 (20.4) | 40 (22.4) | 13 (16.1) | .243 |
| **Smoking (nicotine) status**, Current | 103 (39.6) | 65 (36.3) | 38 (46.9) | .106 |
| **Alcohol Use** |  |  |  |  |
| None | 99 (38.2) | 75 (41.9) | 24 (30.0) | .086 |
| ≤7 drinks/wk | 131 (50.6) | 88 (49.2) | 43 (53.8) |  |
| >7 drinks/wk | 29 (11.2) | 16 (8.9) | 13 (16.2) |  |
| **Marijuana use**, current | 74 (28.5) | 47 (26.3) | 27 (33.3) | .242 |
| **Crack/cocaine/heroin use**, current | 47 (18.1) | 26 (14.5) | 21 (25.9) | **.027** |
| **Menopausal stage** |  |  |  |  |
| Premenopausal/Early<br>perimenopause | 46 (18.1) | 30 (17.2) | 16 (20.0) | .596 |
| Late<br>perimenopause/Postmenopausal | 208 (81.9) | 144 (82.8) | 64 (80.0) |  |
| **BMI**, Median (IQR) | 31.0 (25.0,<br>37.1) | 30.9 (25.0,<br>36.3) | 31.2 (25.2,<br>38.4) | .555 |
| **BMI (obesity)**, ≥30 | 141 (54.7) | 96 (53.9) | 45 (56.3) | .730 |
| **Diabetes**, yes | 53 (21.7) | 39 (23.8) | 14 (17.5) | .264 |
| **Frailty Categories and**<br>**Components (MWCCS)** |  |  |  |  |
| No frailty | 83 (33.7) | 54 (32.0) | 29 (37.7) | .380 |
| Pre-frail /frail | 163 (66.3) | 115 (68.0) | 48 (62.3) |  |
| **Slow (Walk slower than 80<sup>th</sup>**<br>**percentile of overall population)** |  |  |  |  |
| Yes | 103 (40.9) | 72 (41.9) | 31 (38.8) | .640 |
| No | 149 (59.1) | 100 (58.1) | 49 (61.2) |  |
| **Weak (Grip strength test lower**<br>**than 20<sup>th</sup> percentile of overall**<br>**population)** |  |  |  |  |
| Yes | 31 (15.3) | 23 (16.6) | 8 (12.5) | .456 |
| No | 172 (84.7) | 116 (83.4) | 56 (87.5) |  |
| **Physical Limitations** |  |  |  |  |
| Yes | 67 (26.1) | 47 (26.7) | 20 (24.7) | .733 |
| No | 190 (73.9) | 129 (73.3) | 61 (75.3) |  |
| **Exhaustion** |  |  |  |  |
| Yes | 54 (28.6) | 38 (30.4) | 16 (25.0) | .437 |
| No | 135 (71.4) | 87 (69.6) | 48 (75.0) |  |
| **Unintentional weight loss**<br>**(≥10 pounds in past year)** |  |  |  |  |
| Yes | 48 (18.6) | 34 (19.2) | 14 (17.3) | .712 |
| No | 210 (81.4) | 143 (80.8) | 67 (82.7) |  |
| **CESD-10 Depressive Symptoms** |  |  |  |  |
| CES-D ≥ 10 | 61 (24.0) | 41 (23.6) | 20 (25.0) | .803 |
| CES-D <10 | 193 (76.0) | 133 (76.4) | 60 (75.0) |  |
| **Anxiety (CAT-MH)** |  |  |  |  |
| Normal/Mild | 217 (85.8) | 147 (84.5) | 70 (88.6) | .384 |
| Moderate/Severe | 36 (14.2) | 27 (15.5) | 9 (11.4) |  |
| **3-Item Loneliness Scale** |  |  |  |  |
| Loneliness Scale <6 | 187 (71.9) | 130 (72.6) | 57 (70.4) | .708 |
| Loneliness Scale ≥6 | 73 (28.1) | 49 (27.4) | 24 (29.6) | |
| **WWH only (n=179)** |  |  |  |  |
| **HAART, yes** |  | 173 (96.7) |  |  |
| **Time since ART in years, median (IQR)** |  | 17.8 (12.6, 24.3) |  |  |
| **Detectable viral load, >20 copies/ml** |  | 43 (24.0) |  |  |
| **Current CD4/mm3, Median (IQR)** |  | 761 (527, 1061) |  |  |
| **CD4 Count/mm3, >500** |  | 137 (77.0) |  |  |
| Data presented as counts (frequencies), unless otherwise noted.<br>Due to missing data, n's may not add up to total n=260.<br><sup>a</sup>p-value from chi-square tests for categorical variables (Fisher's exact when cells are <5) or t-tests/Mann-Whitney U tests for continuous variables |  |  |  |  |

There were no statistically significant differences in self-reported joint pain by HIV-serostatus when compared as a dichotomous pain vs. no pain variable or when compared as a function of scored pain severity (Table 2). Overall, 34.6% of women reported no joint pain, 23.1% reporting mild, 18.5% reporting moderate, and 23.8% reporting severe joint pain. When asked the site of pain, 60.6% of women reported pain in their knees, 42.4% reported pain in their hips, and 36.5% reported pain in their spine region.

**Table 2.** Joint pain among Chicago Cook County MWCCS women, overall and by HIV serostatus.

|  | **Overall**<br>**(n=260)** | **WWH**<br>**(n=179)** | **WWoH**<br>**(n=81)** | **p-**<br>**value<sup>a</sup>** |
| --- | --- | --- | --- | --- |
| **Summary PEG score\*** (any pain) |  |  |  |  |
| No reported pain | 90 (34.6) | 58 (32.4) | 35 (39.5) | .265 |
| Any reported pain | 170 (65.4) | 121 (67.6) | 49 (60.5) |  |
| **Summary PEG score\*** (pain severity) |  |  |  |  |
| No reported pain | 90 (34.6) | 58 (32.4) | 32 (39.5) | .719 |
| Mild pain | 60 (23.1) | 42 (23.5) | 18 (22.2) |  |
| Moderate pain | 48 (18.5) | 34 (19.0) | 14 (17.3) |  |
| Severe pain | 62 (23.8) | 45 (25.1) | 17 (21.0) |  |
| **PEG domains for those reporting pain (n=170)** |  |  |  |  |
| **Pain on average in the past week** |  |  |  |  |
| Median (IQR) | 7 (4, 9) | 6 (5, 9) | 7 (4, 8) | .730 |
| **Joint pain interfered with enjoyment of life** |  |  |  |  |
| Median (IQR) | 5 (1, 8) | 5 (0, 8) | 5 (1, 7) | .910 |
| **Joint pain interfered with general activity** |  |  |  |  |
| Median (IQR) | 5 (2, 8) | 5 (2, 8) | 5 (1, 7) | .636 |
| **Pain site for those reporting pain (n=170)** |  |  |  |  |
| Hip pain | 72 (42.4) | 50 (41.3) | 22 (44.9) | .669 |
| Knee pain | 103 (60.6) | 74 (61.2) | 29 (59.2) | .812 |
| Spine pain | 62 (36.5) | 46 (38.0) | 16 (32.7) | .511 |
| Other pain | 48 (28.2) | 37 (30.6) | 11 (22.5) | .286 |
| Data presented as counts (frequencies), unless otherwise noted. |  |  |  |  |
| <sup>a</sup>p-value from chi-square tests for categorical variables (Fisher's exact when cells are <5) |  |  |  |  |
| * The summary PEG score was calculated as the average of the three scaled responses. |  |  |  |  |

Women were dichotomized into those reporting no or mild joint pain and compared to those reporting moderate or severe joint pain (Table 3). Women reporting moderate or severe joint pain were more likely to have lower annual household income (defined as <$18,000 annually), less likely to be employed, and more likely to not have a partner. Amongst WWH, those with a longer time since ART initiation were more likely to report no/mild joint pain, although only marginally significant (p=0.059).

**Table 3:** Demographic and lifestyle factors according to PEG-reported pain severity.

|  | No/mild joint pain<br>(n=150) | Moderate/severe joint<br>pain<br>(n=110) | p-value |
| --- | --- | --- | --- |
| **HIV serostatus** |  |  |  |
| WWH | 100 (66.7) | 79 (71.8) | .376 |
| WWoH | 50 (33.3) | 31 (28.2) |  |
| **Age, mean (SD)** | 56.3 (9.0) | 58.2 (7.9) | .083 |
| **Race/Ethnicity** |  |  |  |
| Black | 109 (72.7) | 70 (63.6) | .199 |
| Hispanic | 14 (9.3) | 9 (8.2) |  |
| Other | 20 (13.3) | 26 (23.6) |  |
| White | 7 (4.7) | 5 (4.6) |  |
| **Annual household income, ≤18,000** | 69 (46.3) | 68 (68.0) | **.0007** |
| **Educational Attainment** |  |  |  |
| <High school | 35 (23.3) | 40 (36.4) | .053 |
| High school | 48 (32.0) | 25 (22.7) |  |
| >High school | 67 (44.7) | 45 (40.9) |  |
| **Employed, yes** | 62 (41.6) | 22 (20.2) | **.0003** |
| **Housing stability, lives in own home/apt** | 129 (86.0) | 99 (90.0) | .332 |
| **Partner status, has partner** | 40 (26.7) | 13 (11.8) | **.003** |
| **Smoking (nicotine) status, Current** | 58 (38.7) | 45 (40.9) | .715 |
| **Alcohol Use** |  |  |  |
| None | 51 (34.2) | 48 (43.6) | .291 |
| ≤7 drinks/wk | 81 (54.4) | 50 (45.5) |  |
| >7 drinks/wk | 17 (11.4) | 12 (10.9) |  |
| **Marijuana use, current** | 43 (28.7) | 31 (28.2) | .932 |
| **Crack/cocaine/heroin use, current** | 26 (17.3) | 21 (19.1) | .716 |
| **Menopausal stage** |  |  |  |
| Premenopausal/Early perimenopause | 31 (21.5) | 15 (13.6) | .106 |
| Late perimenopause/Postmenopausal | 113 (78.5) | 95 (86.4) |  |
| **BMI, Median (IQR)** | 30.6 (24.7, 36.5) | 32.1 (25.7, 38.6) | .146 |
| **BMI (Obesity), ≥30** | 76 (50.7) | 65 (60.2) | .130 |
| **WWH only (n=179)** |  |  |  |
| **ART, yes** | 96 (96.0) | 77 (97.5) | .695 |
| **Time since ART initiation (years),**<br>**Median (IQR)** | 19.0 (13.4, 25.0) | 16.7 (10.6, 23.5) | .059 |
| **Detectable viral load, >20 copies** | 24 (24.0) | 19 (24.1) | .994 |
| **Current CD4, Median (IQR)** | 771 (525, 1064.5) | 759 (533, 1032) | .720 |
| **CD4 Count, >500** | 78 (78.0) | 59 (75.6) | .711 |
| Due to missing data, n's may not add up to total n=260. Data presented as counts (frequencies), unless otherwise noted. |  |  |  |
| <sup>a</sup>p-value from chi-square tests for categorical variables (Fisher's exact when cells are <5) or t-tests/Mann-Whitney U tests for continuous variables |  |  |  |

### Relationship between Physical, Mental, and Social Health with Pain Severity

We next compared physical, mental health, and social health by dichotomized joint pain severity in the full study sample (Table 4). There was no effect medication by HIV-serostatus for any of our outcomes of interest (data not shown). Women reporting more severe joint pain were statistically more likely to be pre-frail/frail, based on the Fried Frailty Phenotype (FFP) criteria. We further evaluated each FFP component separately and noted that women reporting more severe joint pain were statistically more likely to walk slower, suffer from exhaustion, and report that their health was more likely to limit their physical activity. Additionally, women reporting more severe joint pain were significantly more likely to report both an increased depressive symptom burden and to have moderate/severe anxiety. Finally, women reporting greater joint pain severity were significantly more likely to report higher levels of loneliness. There were no differences in self-reported unintentional weight loss or in grip strength by joint pain severity.

**Table 4.** Physical, mental, and social health related indices by pain severity.

|  | Overall<br>(n=260) | No/mild PEG<br>score<br>(n=150) | Moderate/severe PEG<br>score<br>(n=110) | p-value |
| --- | --- | --- | --- | --- |
| Physical Health Variables |  |  |  |  |
| Frailty Categories |  |  |  |  |
| No frailty | 83 (33.7) | 64 (43.5) | 19 (19.2) | <.0001 |
| Pre-frail/frail | 163 (66.3) | 83 (56.5) | 80 (80.8) |  |
| Slow (Walk slower than 80<sup>th</sup><br>percentile of overall population) |  |  |  |  |
| No | 149 (59.1) | 98 (66.2) | 51 (49.0) | .006 |
| Yes | 103 (40.9) | 50 (33.8) | 53 (51.0) |  |
| Weak (Grip strength test lower<br>than 20<sup>th</sup> percentile of overall<br>population) |  |  |  |  |
| No | 172 (84.7) | 115 (87.1) | 57 (80.3) | .196 |
| Yes | 31 (15.3) | 17 (12.9) | 14 (19.7) |  |
| Physical Limitations |  |  |  |  |
| No | 190 (73.9) | 137 (91.9) | 53 (49.1) | <.0001 |
| Yes | 67 (26.1) | 12 (8.1) | 55 (50.9) |  |
| Exhaustion |  |  |  |  |
| No | 135 (71.4) | 95 (87.2) | 40 (50.0) | <.0001 |
| Yes | 54 (28.6) | 14 (12.8) | 40 (50.0) |  |
| Unintentional weight loss<br>(≥10 pounds in past year) |  |  |  |  |
| No | 210 (81.4) | 123 (82.5) | 87 (79.8) | .577 |
| Yes | 48 (18.6) | 26 (17.5) | 22 (20.2) |  |
| Mental Health |  |  |  |  |
| CESD-10 Depressive Symptoms |  |  |  |  |
| CES-D <10 | 193 (76.0) | 120 (82.2) | 73 (67.6) | .007 |
| CES-D ≥ 10 | 61 (24.0) | 26 (17.8) | 35 (32.4) |  |
| Anxiety CAT-MH Score |  |  |  |  |
| Normal/Mild (64 or less) | 217 (85.8) | 133 (91.7) | 84 (77.8) | .002 |
| Moderate/Severe (65 or more) | 36 (14.2) | 12 (8.3) | 24 (22.2) |  |
| Social Health |  |  |  |  |
| 3-Item Loneliness Scale |  |  |  |  |
| Loneliness Scale <6 | 187 (71.9) | 115 (76.7) | 72 (65.4) | .047 |
| Loneliness Scale ≥6 | 73 (28.1) | 35 (23.3) | 38 (34.6) |  |
| Due to missing data, n's may not add up to total n=260. Data presented as counts (frequencies), unless otherwise noted. |  |  |  |  |
| a p-value from chi-square tests for categorical variables (Fisher's exact when cells are <5) or t-tests/Mann-Whitney |  |  |  |  |

Stepwise MV models were constructed to evaluate the impact of joint pain on physical, mental, and social health (Table 5). The models were adjusted with demographic variables found to be significantly associated with each of the four outcome variables (frailty, depressive symptoms, anxiety, and loneliness) presented in Supplemental Table 2). In the full study sample, women with moderate/severe joint pain were 3 times more likely to be pre-frail/frail (AOR 3.05), after adjusting for age, HIV-serostatus and smoking. The results were similar in models with WWH only, with moderate/severe joint pain associated with a greater likelihood of being classified as pre-frail/frail (AOR 2.35), after adjusting for age, smoking and time since ART initiation.

**Table 5:**
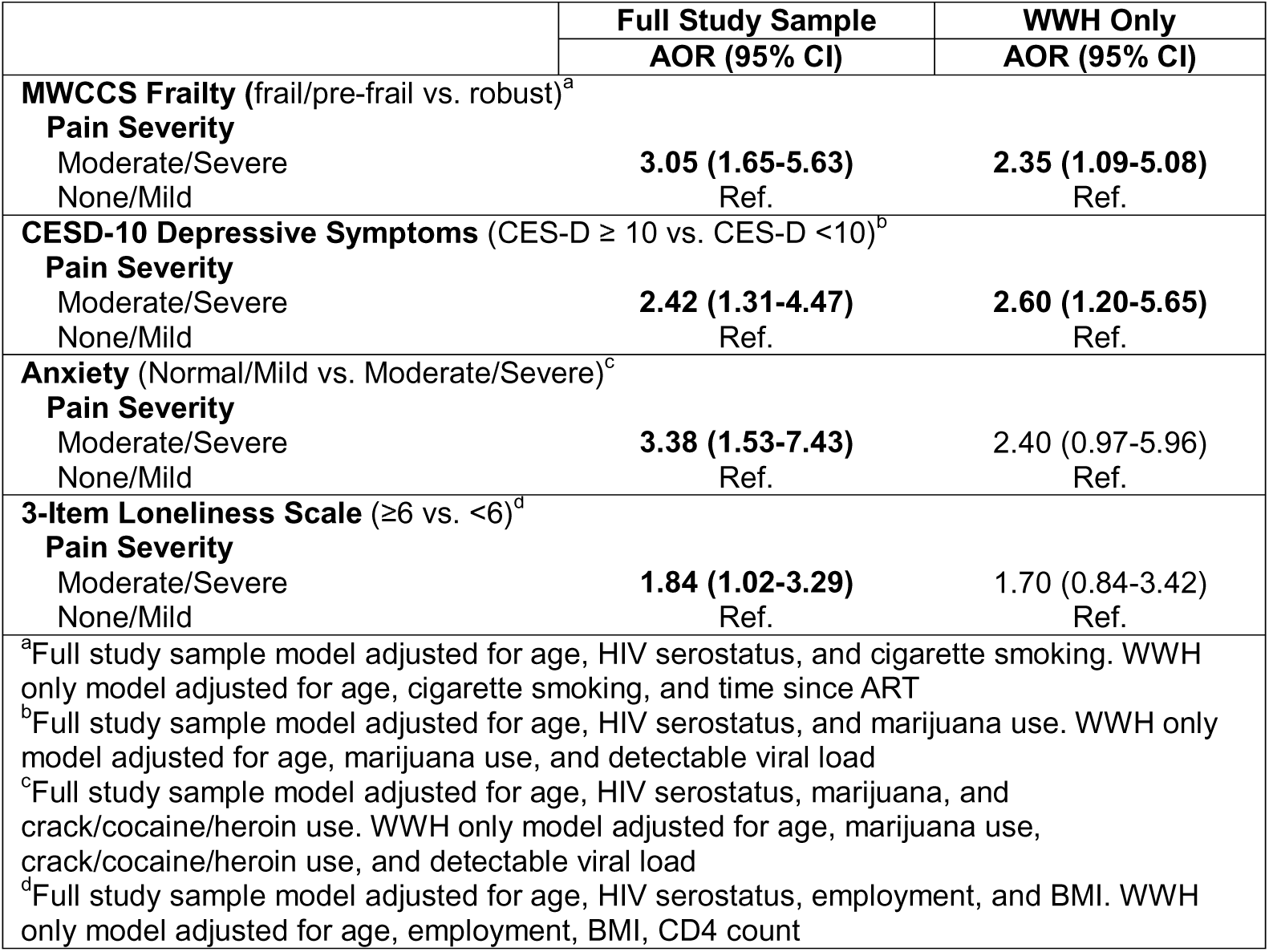
Relationship between self-reported joint pain and physical and mental health.

Moderate/severe joint pain also statistically significantly increased the likelihood for depressive symptoms, both in the full study sample (AOR 2.42) after adjusting for age, HIV-serostatus and marijuana use and in WWH only (AOR 2.60) after adjusting for age, marijuana use and viremia. Similar results were noted with anxiety as the outcome. In the full study sample, moderate/severe joint pain statistically significantly increased the likelihood of anxiety (AOR 3.38) after adjusting for age, HIV-serostatus, marijuana and crack, cocaine, and heroin use however in WWH only, the results were no longer statistically significant (AOR 2.40) and were qualitatively decreased compared to the full study sample.

Moderate/severe joint pain increased the likelihood of loneliness. In models including the full study sample, the relationship was statistically significant with an AOR of 1.84 after adjusting for age, HIV-serostatus, employment and BMI. However, in models limited to WWH, the relationship was no longer statistically significant, despite being qualitatively similar, with an AOR of 1.70.

## Discussion

Chronic joint pain is a growing public health problem. Painful joint conditions, such as osteoarthritis (19) and chronic low back pain (20), are becoming more common as the population ages. Importantly, these conditions are associated with substantial morbidity and healthcare costs (21). People with HIV (PWH) now experience a lifespan similar to those without HIV (2), therefore the number of PWH suffering from joint pain and related conditions is likely to increase. Further, HIV is a conditioned characterized by chronic low-grade inflammation, which is thought to increase the burden of age-related musculoskeletal conditions in PWH (22). Previous studies evaluating the prevalence of self-reported joint or musculoskeletal pain in PWH have noted prevalence ranges between 25 and 69% but did not include comparisons to similar populations of persons without HIV (2, 23, 24). The current study reports the prevalence and severity of self-reported joint pain in midlife women with and without HIV enrolled in a longitudinal cohort study and represents a population at high risk for joint pain due to their age, menopause status, and high BMI (3, 4). The lack of statistically significant differences between WWH and WWoH may be partly due to the relatively small sample size of our study sample but also indicates that factors other than HIV likely have a greater impact on joint pain. Although interestingly, we did detect a near significant trend for years on ART, with WWH with the longest ART treatment duration having have a slightly lower prevalence of moderate/severe joint pain. It will be interesting to determine whether this effect becomes significant in larger samples sizes and whether the relationship between joint pain and ART use is influenced by the specific ART formulations used.

Our study found that women with greater joint pain were statistically significantly more likely to be classified as pre-frail or frail as opposed to a robust physical phenotype. However, this association was not impacted by HIV-serostatus. Prior to the onset of widespread ART availability and use, PWH were noted to have greater risk for frailty (25). Recent studies suggest that frailty prevalence in WWH is no longer significantly greater than demographically similar WWoH (26), which we confirm in the current study. However, the adverse effects of frailty persist despite improved ART (27), with increased risks for falls (28) and fractures (29). While the current study modeled joint pain as the predictor, it is worth acknowledging that frailty has been reported to worsen joint pain (30). Therefore, a bidirectional relationship likely exists between joint pain and frailty that we plan to assess in follow-up studies.

Our results also show a statistically significant association between joint pain and greater anxiety and depressive symptoms, which is consistent with data reported in the larger studies of people without HIV (31). Prior studies also indicate that the presence of depressive or anxiety symptoms reduces treatment adherence for painful musculoskeletal conditions, such as rheumatoid arthritis, which may worsen pain (32, 33) and contribute to the bidirectional relationship reported between joint pain and depression (34). Interestingly, in our study, the associations between joint pain and anxiety were no longer statistically significant in models with WWH only. We hypothesize that this loss of statistical significance is attributable to small sample size as there are relatively few WWH (n=27) in our study with moderate/severe anxiety.

Women with greater joint pain were more likely to experience loneliness. Previous studies have confirmed the influence of chronic pain (37) and musculoskeletal pain specifically on loneliness (11). In longitudinal studies, loneliness is more common years before the onset of chronic pain and only gets worse after first reporting of pain (38). The cause may be the negative impact that severe joint pain has on mobility and independence. Further, joint pain may also influence social relationships, as we noted that women with more moderate/severe joint pain were less likely to be partnered. Surprisingly, the association between joint pain severity and loneliness was no longer significant in WWH. While it is possible that this may be attributed to greater social support networks for WWH, it is worth noting that there were no differences in the self-reported loneliness according to HIV-serostatus.

To our knowledge, this is the first study to investigate the prevalence, severity and potential impact of joint pain among women with HIV and a comparison group of women without HIV. Importantly, our study takes the first step towards addressing joint pain in people with HIV, as suggested by the Global Task Force for Chronic Pain in PWH (39), and by patients themselves (40). Our study benefited from a well-matched population of mid-life women with HIV and women without HIV and the results suggest that WWH are not at a greater risk for joint pain, nor do they suffer from significantly greater joint pain severity than demographically similar WWoH. However, the prevalence of any joint pain was high (65%), with over 40% of our cohort reporting moderate/severe joint pain. Our study does confirm that joint pain in WWH contributes to worse physical, mental and social health, stressing the importance of targeted joint pain interventions or treatments. Importantly, there are few studies that have investigated treatment for chronic pain in people with HIV and most have been low quality, with limited sample sizes, short follow-up periods, and few randomization studies (41). Therefore, future studies should seek to evaluate the efficacy of conventional joint pain treatment strategies in WWH, identify novel and/or adjuvant therapies, and determine whether alleviating joint pain can improve physical, mental and/or social function.

The current study has several limitations. The relatively small sample size limited our ability to detect specific HIV effects. We focused on midlife women and therefore cannot address sex differences, and we conducted our study at a single site in Chicago, so cannot address potential geographic differences in joint pain prevalence or correlates. We did not have access to joint disease diagnoses such as rheumatoid, osteoarthritis, history of septic arthritis or joint injury, therefore we could not determine the contributing role of these conditions to joint pain severity or functional impairment. Finally, our study was a cross-sectional evaluation of the correlates of joint pain and as noted above, bidirectional relationships have been reported for our outcomes of interest – frailty, depression, anxiety and loneliness. Longitudinal assessment of joint pain in the full MWCCS will allow us to not only determine the prevalence and severity of joint pain in men and women by anatomical location across various age groups, geographical locations, and by HIV status but also allow us to examine temporal relationships between joint pain progression, functional outcomes, and chronic comorbidities.

## Conclusions

Joint pain is highly prevalent amongst mid-life women with and without HIV. More severe joint pain is associated with significantly increased likelihood of being pre-frail or frail and endorsing increased anxiety and depressive symptoms, and loneliness. Efforts to mitigate joint pain have the potential to contribute to improved physical, mental, and social health in women with HIV and demographically similar women without HIV.

## Data Availability

All data produced in the present study are available upon reasonable request to the MWCCS

## Author Contributions

All authors contributed to the study and met the criteria for authorship. Conceptualization: RDR and KMW. Data curation and formal analysis: ED. Funding Acquisition: RDR, ALF, MHC. Investigation: ED, RH, DJ. Methodology: RDR, ED, SMC, TEW, ALF, MHC, KMW. Project Administration: RDR, ED, TEW, ALF, MHC, KMW. Supervision: RDR, ED, TEW, ALF, MHC, KMW. Writing – Original Draft Preparation: RDR, SMC, KMW. Writing – Review & Editing: RDR, ED, SMC, TEW, ALF, MHC, KMW. RDR, SMC, and KMW

## Acknowledgements

Data in this manuscript were collected by the MACS/WIHS Combined Cohort Study (MWCCS). We would like to first acknowledge the contributions of the women enrolled in the Chicago Cook County Clinical Research Site of the MWCCS who participated in this study.

## Data Availability

The data that supports the findings of this study are available on request from the MWCCS. That data are not publicly available due to privacy or ethical restrictions. Interested researchers can request a deidentified dataset from the MWCCS Data Analysis and Coordination Center (DACC).

